# Eosinopenia Phenotype in Patients with Coronavirus Disease 2019: A Multi-center Retrospective Study from Anhui, China

**DOI:** 10.1101/2020.04.22.20071050

**Authors:** Yusheng Cheng, Yun Zhou, Mengde Zhu, Lei Zha, Zhiwei Lu, Zhen Ding, Yang Jianghua, Gang Yang

## Abstract

**Background:** Coronavirus disease 19 (COVID-19) has become a global unprecedented pandemic infecting more than one millon people, which is declared by WHO as a international public health emergency. Eosinopenia may predict a poor prognosis of COVID-19. However, to date, there is no detailed analysis of the clinical characteristics of COVID-19 patients with eosinopenia.

**Research question:** The aim of this study was to describe clinical characteristics of COVID-19 patients with eosinopenia.

**Study Design and Methods:** This was a multi-center retrospective study conducted in three tertiary hospitals. A total of 59 patients with COVID-19 were reviewed from January 23, 2020 to March 10, 2020. We described clincial characteristics of patients with COIVD-19 and eosinopenia phenotype.

**Results:** The median age of patients with COVID-19 was 39 years old, and 32 (54,2%) were male. Patients with severe type had higher proportions of dyspnea (50%) and gastrointestinal symptoms (50%) compared with mild or moderate patients. Laboratory findings indicated that lower counts of lymphocyte and eosnophils were observed in patients with severe type. Cough, sputum, and fatigue were more common symptoms in eosinopenia patients compared with non-eosinopenia patients. High proportion of comorbidities was observed in eosinopenia patients. Laboratory findings indicated that lymphocyte counts (median: 101 cells/μl) in eosinopenia patients were significantly less than those of non-eosinopenia patients (median: 167 cells/μl, *p*<0.001). The use of corticosteroids therapy in COVID-19 patients with eosinopenia were notably higher than those in patients with non-eosinopenia (50% vs 13.8%, respectively, *p*=0.005). Compared with parameters in non-eosinopenia patients, eosinopenia patients were more inclined to have less lymphocyte counts (OR value 6.566, 95%CI[1.101-39.173], *p*=0.039).

**Interpretation:** Eosinopenia are very common in COVID-19 patient, particularly in severe patients. Common symptoms included fever, cough, sputum, and fatigue are frequent in eosinopenia patients. Eosinopenia may represent a novel phenotype in COVID-19, which needs further investigation.

## Introduction

Since December, 2019, a series of resembling viral pneumonia cases of unknown cause have occured in Wuhan, China, with clinical presentations inclued fever, cough, and myalgia or fatigue, dyspnoea, sputum production, headache, haemoptysis, and diarrhoea. This resembling viral pneumonia was similar to the severe respiratory illness caused by severe acute respiratory syndrome coronavirus^1^. Lower respiratory tract samples from patients with novel coronavirus pneumonia (NCP) analyzed by sequencing indicated a novel coronavirus named 2019 novel coronavirus (2019-nCoV) ^2^. Chan JF and colleagues first provided data from phylogenetic analysis of genetic sequences indicated person-to-person transmission of 2019-nCoV ^3^. On February 11, 2020, the World Health Organization (WHO) officially named NCP as Coronavirus Disease 2019 (COVID-19), and 2019-nCoV was named severe acute respiratory syndrome coronavirus 2 (SARS-Cov-2) by the International Committee on Taxonomy of Viruses (ICTV).

COVID-19 has become a global unprecedented pandemic infecting more than one millon people, which is declared by WHO as a international public health emergency. The overall rate of severe cases was 16.0%, and mortality was 3.2% in a nationwide analysis of China^4^. Elderly people, complicated with comorbidities, impaired immune function, and involvement of multiple lung lobes are risk factors of patients for having severe or critical disease^5,6^. At an early stage, increased serum d-dimer predicts a poor prognosis pf COVID-19 patients^7^. Meanwhile, lower counts of CD3^+^CD8^+^ T cells and high cardiac troponin I are two predictors for high mortality of COVID-19^8^.

Dysregulations of immune response are features of COVID-19, such as decreased immune cell counts and elevated inflammatory cytokines^9^. Mounting evidence has shown that lymphopenia is common in patients with COVID-19, which is close related to severity of COVID-19^10-12^. Of note, Du Y and colleagues found that 81.2% fatal cases had very low counts of blood eosinophil at admisson, and eosinopenia predicted a poor prognosis of COVID-19^13^. Normalization of blood eosinophil after treatment may be an indicator of COVID-19 improvement^14^. However, to date, there is lack of detailed analysis of COVID-19 patients with eosinopenia. Here, the aim of this study was to describe clinical characteristics of COVID-19 patients with eosinopenia phenotype in order to improve the knowledge of COVID-19.

## Methods

### Study design and participants

This was a multi-center retrospective study conducted in three tertiary hospitals: Yijishan hosptial of Wanan medical college, the second people’s hospital of Wuhu city, and the first people’s hospital of Hefei city. All COVID-19 patients were reviewed from January 23, 2020 to March 10, 2020. This study was approved by the Ethics Committees of Yijishan hosptial of Wanan medical college,the second people’s hospital of Wuhu city, and the first people’s hospital of Hefei city (No.20200101). The informed consent from each COVID-19 patient was waived since this study followed the the policy for public-health-outbreak investigation of emerging infectious diseases issued by the National Health Commission of the People’s Republic of China.

### Data collection

From electronic medical records, data collected included demographics, underlying diseases, medical history, comorbidities (hypertension, coronary heart disease or diabetes mellitus, etc), symtoms (highest temperature, cough, sputum, dyspnea, fatigue, myalgia, and gastrointestinal symptoms), signs, laboratory tests, chest CT scans, clinical treatment (i.e. corticosteroid therapy, antiviral therapy, convalescent plasma therapy, and tocilizumab therapy, etc). All COVID-19 patients were diagnosed based on clinical presentations, chest imaging features, and SARS-COV-2 detected in respiratory tract samples. According to the fifth version of the guidelines for diagnosis and Treatment of COVID-19 issued by the National Health Commission of China, patients were divided into four types: mild, moderate, severe, and critical type.

### Statistical analysis

Categorical variables were reported using numbers and percentages, and continuous variables were presented as the medians (interquartile ranges[IQRs]). The Kruskal-Wallis H test or Mann-Whitney U test was used for continuous data. For categorical data, the χ2 test was used. The multivariate logistic regression analysis was used to differentiate independent risk factors associated with eosinopenia phenotype in COVID-19 patients. The *p* value less than 0.05 was considered to be statistically significant. The SPSS 22.0 software (IBM Corp., Armonk, NY) was used for data analysis.

## Results

### Baseline clinical characteristics of patients with COVID-19

In this multi-center retrospective study, a total of 59 patients with COVID-19 were reviewed from January 23, 2020 to March 10, 2020. Of the 59 patients, 5 were mild, 46 were moderate, 8 were severe. The median age of patients with COVID-19 was 39 years old, and 32 (54,2%) were male. Patients with severe type was significantly older than patients with mild or moderate type (median age: 57 vs 24 vs 37 years old, p=0.001). Patients with severe type had higher proportions of dyspnea (50%) and gastrointestinal symptoms (50%) than patients with mild to moderate type (*p*<0.05) (Table 1). Other common symptoms included cough, sputum, fatigue, and myalgia did not reach statistic differences among these types. Fifteen of 59 patients (25.4%) had one or more comorbidities, mostly in patients with severe type (62.5%, *p*=0.020) (Table 1). Laboratory findings indicated that increases of D-Dimer (median concentration: 1ng/ml) and CRP (median concentration: 88.2 mg/dl) were observed in patients with severe type compared with mild to moderate type (median concentration: 0.19 ng/ml and 0.5 ng/ml, *p* =0.01; 0.86 mg/dl and 12.64 mg/dl, *p*=0.007; respectively). Lower counts of lymphocyte (median counts: 900 cells/ μ l) and eosnophils (median counts: 100 cells/μl) were observed in patients with severe type, though there were no statistic differences among these types (Table 1). All patients received CT scans, radiological features of lungs showed that 5 of patients (8.5%) were normal, unilateral pneumonia occurred in 11 patients (18.5%), and 43 patients (72.9%) had bilateral pneumonia (Table 1).

**Table 1.** Baseline clinical characteristics of patients with COVID-19

|  | Total (n=59) | Mild (n=5) | Moderate (n=46) | Severe (n=8) | *p* |
| --- | --- | --- | --- | --- | --- |
| Age (years) | 39.0 (30.0, 54.0) | 24 (12.5, 33.5) | 37 (30.8, 53.0) | 57.0 (51.75, 66.5) | 0.001 |
| Male (%) | 32 (54.2) | 3 (60.0) | 25 (54.3) | 4 (50.0) | 0.939 |
| Highest temperature (°C) | 38 (37.4, 38.4) | 37.7 (37.5, 38.7) | 38 (37.4, 38.4) | 37.85 (37.08, 38.38) | 0.802 |
| Cough | 37 (62.7) | 3 (60.0) | 26 (56.5) | 8 (100.0) | 0.063 |
| Sputum | 18 (30.5) | 2 (40.0) | 11 (23.9) | 5 (62.5) | 0.081 |
| Dyspnea | 9 (15.3) | 0 (0) | 5 (10.9) | 4 (50.0) | 0.011 |
| Fatigue | 30 (50.8) | 3 (60.0) | 21 (45.7) | 6 (75.0) | 0.282 |
| Myalgia | 12 (20.3) | 0 (0) | 11 (23.9) | 1 (12.5) | 0.379 |
| Gastrointestinal symptoms | 11 (18.6) | 1 (20.0) | 6 (13.0) | 4 (50.0) | 0.046 |
| Comorbidities | 15 (25.4) | 0 (0) | 10 (21.7) | 5 (62.5) | 0.020 |
| Laboratory examinations |  |  |  |  |  |
| White blood cell (×10<sup>9</sup>/L) | 5.0 (4.06, 6.5) | 4.92 (3.55, 9.47) | 4.89 (3.92, 6.03) | 6.96 (4.85, 9.6) | 0.095 |
| Lymphocyte (×10<sup>9</sup>/L) | 1.27 (0.9, 1.7) | 1.21 (1.05, 2.98) | 1.35 (0.93, 1.72) | 0.9 (0.45, 1.31) | 0.053 |
| Eosnophils (×10<sup>9</sup>/L) | 0.01 (0.01, 0.05) | 0.02 (0.01, 0.26) | 0.02 (0.00, 0.05) | 0.01 (0.01, 0.33) | 0.384 |
| Eosinopenia | 30 (50.8) | 2 (40) | 22 (47.8) | 6 (75) | 0.321 |
| Haemoglobin (g/L) | 132.0 (120.0, 146.0) | 132 (118.5, 142.5) | 135.5 (121.0, 149.0) | 123.5 (118.25, 133.5) | 0.342 |
| Platelet (×10<sup>9</sup>/L) | 164.0 (138.0, 204.0) | 270 (176.5, 329.0) | 158.5 (131.0, 191.5) | 193.5 (169.0, 207.0) | 0.023 |
| D-Dimer (ng/ml) | 0.5 (0.19, 0.91) | 0.19 (0.14, 0.38) | 0.5 (0.20, 0.78) | 1.00 (0.61, 1.52) | 0.010 |
| ALT (U/L) | 27.0 (17.0, 40.0) | 24.0 (10.5, 31.5) | 26.5 (17.0, 40.0) | 41.0 (21.0, 69.5) | 0.184 |
| AST (U/L) | 23.0 (20.0, 32.0) | 24.0 (21.5, 27.0) | 23.0 (19.0, 29.25) | 38.5 (21.25, 70.25) | 0.127 |
| C-reactive protein (mg/dl) | 15.96 (4.76, 55.15) | 0.86 (0.5, 21.6) | 12.64 (4.76, 39.46) | 88.2 (31.79, 124.68) | 0.007 |
| Radiological features of lung |  |  |  |  |  |
| Normal | 5 (8.5) | 5 (100.0) | 0 (0) | 0 (0) | - |
| Unilateral pneumonia | 11 (18.6) | 0 (0) | 10 (21.7) | 1 (12.5) | - |
| Bilateral pneumonia | 43 (72.9) | 0 (0) | 36 (78.3) | 7 (87.5) | - |

Abbreviations: COVID-19, Corona Virus Disease 2019; SPO<sub>2</sub>, peripheral capillary oxygen saturation; ALT, alanine transaminase; AST, aspartate aminotransferase

### Treatments and prognosis of patients with COVID-19

Nineteen of 59 patients (32.2%) were treated with corticosteroids after admisson. The use of corticosteroids therapy was highest in severe COVID-19 patients (75%) than in mild (0) and moderate (28.3%) patients (*p*=0.009) (Table 2). All patients received antiviral treatments including inhalation of recombinant interferon, lopinavir/ritonavir, arbidol, and oseltamivir. One severe patient received convalescent plasma therapy and tocilizumab (Table 2). Radiographic improvement time was longer in patients with severe type than that in moderate type (median days: 10.5 vs 8.5, respectively, *p*=0.001). No statistic differences were observed in viroloy improvement time and hospital stay among mild to severe type (Table 2).

**Table 2.** Treatments and prognosis of patients with COVID-19

| Table 2. Treatments and prognosis of patients with COVID-19 |  |  |  |  |  |
| --- | --- | --- | --- | --- | --- |
|  | Total (n=59) | Mild (n=5) | Moderate (n=46) | Severe (n=8) | *p* |
| Corticosteroids therapy | 19 (32.2) | 0 (0) | 13 (28.3) | 6 (75.0) | 0.009 |
| Antiviral treatments |  |  |  |  |  |
| Recombinant interferon-α2b | 57 (96.6) | 5 (100) | 45 (97.8) | 7 (87.5) | 0.300 |
| Lopinavir/ritonavir | 57 (96.9) | 5 (100) | 46 (100.0) | 6 (75.0) | 0.001 |
| Arbidol | 26 (44.1) | 0 (0) | 23 (50.0) | 3 (37.5) | 0.094 |
| Oseltamivir | 10 (16.9) | 1(20.0) | 6 (13.0) | 3 (37.5) | 0.231 |
| Convalescent plasma therapy | 1 (1.6) | 0 | 0 | 1 (12.5) | - |
| Tocilizumab | 1 (1.6) | 0 | 0 | 1 (12.5) | - |
| Radiographic improvement time (days) | 8.0 (5.0, 12) | 0 | 8.5 (6.0, 11.25) | 10.5 (5.75, 14.75) | 0.001 |
| Virology improvement time (days) | 11.0 (7.0, 14.0) | 8.0 (4.5, 22.5) | 11.50 (7.75, 13.25) | 5.5 (5.0, 15.25) | 0.624 |
| Hospital stay (days) | 16.0 (11.0, 19.0) | 12.0 (10.0, 22.0) | 16.0 (12.0, 19.0) | 17.0 (10.25, 21.25) | 0.923 |
| Abbreviations: COVID-19, Corona Virus Disease 2019 |  |  |  |  |  |

### Clincial characteristics of eosinopenia phenotype in COVID-19 patients

The median age of COVID-19 patients with eosinopenia was significantly higher than that in patients with non-eosinopenia (47 vs 36, respectively, *p*=0.042). Eosinopenia patients had higher temperature than non-eosinopenia patients (median highest temperature: 38 vs 37.7, *p*=0.02). Cough, sputum, and fatigue were more common symptoms in eosinopenia patients compared with non-eosinopenia patients (80% vs 44.8, *p*=0.007; 46.7% vs 13.8, *p*=0.01; 63.3% vs 37.9, *p*=0.07; respectively). High proportion of comorbidities (33.3%) was observed in eosinopenia patients, which was 17.2% in non-eosinopenia patients. Eosinopenia patients had lower SpO2 than non-eosinopenia patients (median value: 97 vs 98, *p*=0.039) (Table 3). Laboratory findings indicated that lymphocyte counts (median: 101 cells/μl) in eosinopenia patients were significantly less than those of non-eosinopenia patients (median: 167 cells/μl, *p*<0.001). The AST concentrations (median value: 28.5 U/L) in eosinopenia patients were higher than those of non-eosinopenia patients (median value: 23 U/L, *p*=0.048). Other laboratory parameters as well as radiological features of lung did not reach statistic differences between the two groups (Table 3).

**Table 3.** Comparsion of characteristics of eosinopenia and non-eosinopenia phenotypes in COVID-19 pat

|  | non-Eosinopenia (n=29) | Eosinopenia (n=30) | *p* |
| --- | --- | --- | --- |
| Age (years) | 36.0 (26.0, 52.5) | 47 (35, 56.25) | 0.042 |
| Male (%) | 17 (58.6) | 15 (50) | 0.604 |
| Highest temperature (°C) | 37.7 (37.3, 38.0) | 38 (37.72, 38.5) | 0.02 |
| Cough | 13 (44.8) | 24 (80) | 0.007 |
| Sputum | 4 (13.8) | 14 (46.7) | 0.01 |
| Dyspnea | 3 (10.3) | 6 (20.0) | 0.472 |
| Fatigue | 11 (37.9) | 19 (63.3) | 0.07 |
| Myalgia | 6 (20.7) | 6 (20.0) | 1.00 |
| Comorbidities | 5 (17.2) | 10 (33.3) | 0.233 |
| SpO2 (%) | 98 (97, 98.5) | 97 (94.5, 98.0) | 0.039 |
| Gastrointestinal symptoms | 4 (13.8) | 7 (23.3) | 0.506 |
| Laboratory examinations |  |  |  |
| White blood cell (×10<sup>9</sup>/L) | 5.26 (4.59, 6.68) | 4.46 (3.81, 6.06) | 0.124 |
| Lymphocyte (×10<sup>9</sup>/L) | 1.67 (1.22, 2.04) | 1.01 (0.68, 1.38) | < 0.001 |
| Haemoglobin (g/L) | 132.0 (122.5, 149.0) | 131.5 (117.25, 144.5) | 0.367 |
| Platelet (×10<sup>9</sup>/L) | 165.0 (141.0, 252.0) | 160.0 (131.0, 191.5) | 0.08 |
| D-Dimer (ng/ml) | 0.5 (0.23, 0.71) | 0.58 (0.19, 0.98) | 0.499 |
| ALT (U/L) | 27.0 (17.0, 39.0) | 29.5 (16.75, 43.0) | 0.808 |
| AST (U/L) | 23.0 (18.5, 26.5) | 28.5 (21.75, 35.25) | 0.048 |
| C-reactive protein (mg/dl) | 12.75 (3.49, 37.5) | 22.37 (5.05, 115.03) | 0.178 |
| Radiological features of lung |  |  | 0.862 |
| Normal | 3 (10.3) | 2 (6.7) |  |
| Unilateral pneumonia | 5 (17.2) | 6 (20) |  |
| Bilateral pneumonia | 21 (72.4) | 22 (73.3) |  |

Abbreviations: COVID-19, Corona Virus Disease 2019; SPO2, peripheral capillary oxygen saturation; ALT, alanine transaminase; AST, aspartate aminotransferase

### Treatments and prognosis of COVID-19 patients with eosinopenia

After admission, the use of corticosteroids therapy in COVID-19 patients with eosinopenia were notably higher than those in patients with non-eosinopenia (50% vs 13.8%, respectively, *p*=0.005). Radiographic improvement time (median days: 9.5) and hospital stay (median days: 17) in patients with eosinopenia were numerically longer than those in patients with non-eosinopenia (median days: 6.0 and 15, respectively) (Table 4). However, there were not statistic differences between the two groups. All patients received inhalation of recombinant interferon. Twenty-eight of 30 eosinopenia patients were treated with lopinavir/ritonavir, 14 with arbidol, and 6 with oseltamivir. Convalescent plasma therapy and tocilizumab were used in one severe patient with eosinopenia (Table 4).

**Table 4.** Treatments and prognosis of eosinopenia and non-eosinopenia phenotypes in COVID-19 patients

|  | non-Eosinopenia (n=29) | Eosinopenia (n=30) | *p* |
| --- | --- | --- | --- |
| Corticosteroids therapy | 4 (13.8) | 15 (50) | 0.005 |
| Antiviral treatments |  |  |  |
| Recombinant interferon-α2b | 28 (100) | 29 (100) | - |
| Lopinavir/ritonavir | 29 (100) | 28 (93.3) | 0.492 |
| Arbidol | 12 (41.4) | 14 (46.7) | 0.795 |
| Oseltamivir | 4 (13.8) | 6 (20.0) | 0.731 |
| Convalescent plasma therapy | 0 (0.00) | 1 (3.33%) | - |
| Tocilizumab | 0 (0.00) | 1 (3.33%) | - |
| Radiographic improvement time (days) | 6.0 (3.0, 12.0) | 9.5 (6.75, 11.25) | 0.073 |
| Virology improvement time (days) | 11.0 (7.0, 14.0) | 11.5 (5.75, 14.0) | 0.921 |
| Hospital stay (days) | 15.0 (10.5, 19.0) | 17.0 (12.0, 19.0) | 0.305 |

Abbreviations: COVID-19, Corona Virus Disease 2019

**Table 5.** Multivariate analysis of independent risk factors for differentiating eosinopenia phenotype in COVID-19 patients

|  | OR | 95% CI | *p* |
| --- | --- | --- | --- |
| Age | 0.975 | 0.927-1.025 | 0.314 |
| Highest temperature (°C) | 0.294 | 0.081-1.070 | 0.063 |
| Cough | 1.022 | 0.182-5.514 | 0.980 |
| Sputum | 0.254 | 0.037-1.758 | 0.165 |
| SpO2 | 1.069 | 0.823-1.390 | 0.617 |
| Lymphocyte counts | 6.566 | 1.101-39.173 | 0.039 |
| AST | 0.966 | 0.893-1.044 | 0.38 |
| Corticosteroids therapy | 0.62 | 0.130-2.950 | 0.548 |
| Abbreviations: COVID-19, Corona Virus Disease 2019; SpO2, peripheral capillary oxygen saturation; AST, aspartate aminotransferase |  |  |  |

### Multivariate analysis in COVID-19 patients

Comparsion of characteristics, treatments, and prognosis of eosinopenia patients and non-eosinopenia patients, all variables with a p value blow 0.05 in the univariate analysis were entered into multivariate logistic regression analysis. Compared with parameters in non-eosinopenia patients, eosinopenia patients were more inclined to have less lymphocyte counts (OR value 6.566, 95%CI[1.101-39.173], *p*=0.039).

### Eosnophils in convalescent COVID-19 patients with severe type

In this study, all COVID-19 patients with severe type were cured. The counts of eosnophils as well as lymphocyte were dramatically elevated and normalized in recovery patients with severe type compared with those at adimission. The counts of white blood cells did not differ between both groups (Figure 1).

**Figure 1.**
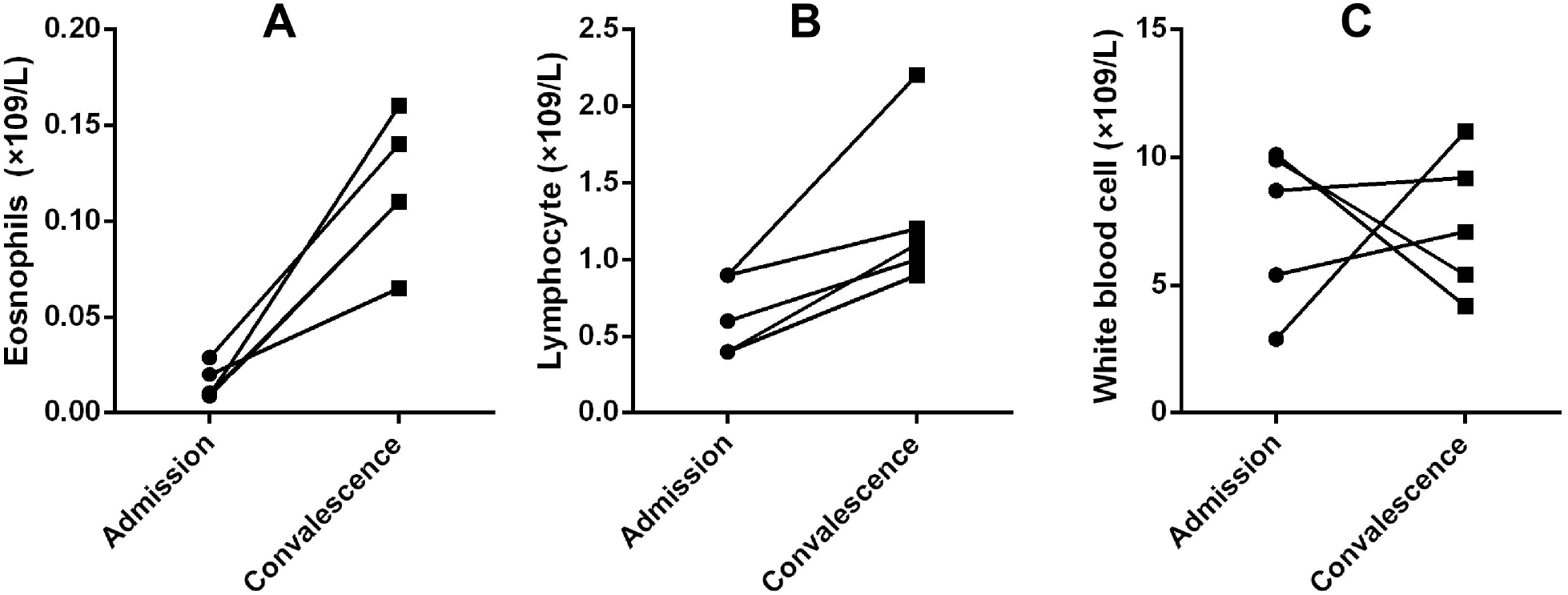
Comparison of white blood cells in COVID-19 patients at adimission and discharge. (A) The counts of eosnophils in COVID-19 patients at adimission and discharge (B) The counts of lymphocyte in COVID-19 patients at adimission and discharge (C) The counts of whole white blood cells in COVID-19 patients at adimission and discharge.

## Disscussion

COVID-19 has become an emerging, and rapidly evolving public health emergency worldwide. By unbiased sequencing from samples from patients, a novel virus named SARS-Cov-2 (previously named 2019-nCoV) was confirmed to be the seventh meber of the coronaviruses family^2^. Recent meta-analysis of data from patients with COVID-19 indicates that fever, cough, myalgia or fatigue, expectoration, and dyspnea are main clinical symptoms. Other symptoms included headache, diarrhea, nausea and vomiting are minor. Remarkably, lymphopenia is frequent in COVID-19 patients^1,15,16^. Tao Chen and colleagues reported that higher proportion of lymphopenia in deceased patients with COVID-19 than that in recovered patients, implicating that lymphopenia might be a predictor of poor outcome^11^. In the present study, we reported clinical characteristics of patients with COVID-19 outside of Hubei province, and first described the clinical features of a novel type of COVID-19: eosinopenia phenotype. It is hoped that results of this study will deep the knowledge of COVID-19 in the future work.

Adequate evaluation of the severity in COVID-19 is of crucial importance. Mounting evidence have indicated that eldly COVID-19 patients are inclined to develop severe condition, and higher rates of dyspnea, lymphopenia, CRP, and D-dimer in severe patients than those in moderate patients^10,11,13,17^. Consistent with previous studies, we found that the median age of COVID-19 patients with severe type were significantly older than patients with mild or moderate type, and severe patients had higher proportions of dyspnea and gastrointestinal symptoms than patients with mild to moderate type. Meanwhile, the results of this study indicated that the proportions of cough, sputum, fatigue, and myalgia were numerically high in severe patients. Chest CT plays an vital role in differential diagnosis of COVID-19. Li X, etal reported that around 83% COVID-19 patients had more than two lobes involved, and most images were multiple lesions localized in the peripheral of bilateral lungs^18^. In this study, all patients received chest CT scans, the radiological features of lungs showed that 72.9% COVID-19 patients and 87.5% severe patients had bilateral pneumonia. We found that one-third of COVID-19 patients were treated with corticosteroids which was highest in severe patients (75%). All patients received antiviral treatments, and one severe patient received convalescent plasma therapy and tocilizumab. No statistic differences were observed in hospital stay among patients in this study may due to lack of adequate knowledge of this unprecedented pandemic.

In the present study, lymphopenia, and increases of D-dimer and CRP were more common in severe COVID-19 patients. SARS-CoV-2 may primarily infect T lymphocytes, resulting in lymphopenia as well as decrease of IFN-γ production^10^. Lymphopenia and elevated CRP may be useful predictors for developments of pneumonia and acute respiratory distress syndrome (ARDS) in COVID-19 patients^19^. Dysfunctions of blood coagulation in severe COVID-19 patients should be paid more concerns. The prevalence of venous thromboembolism (VTE) in COVID-19 patients is around 25 %. D-dimer is a promising biomarker for identifying high-risk groups of VTE^20^. Of note, in this study, more than half of COVID-19 patients developed eosinopenia, particularly severe patients (75%). Similar results were reported in previous studies^9,13,14^. Eosinopenia may indicate a poor prognosis of COVID-19 patients. Autopsy of deceased COVID-19 revealed minimal eosinophils or eosinophils in lung tissues^21,22^. However, the pathogenesis of eosinopenia in COVID-19 remains to be determined, which may be related to depletion of CD8 T cells producing IL-5 to stimulate eosinophil proliferation, or eosinophil consumption caused by higher viral load of SARS-CoV-2^13,14^.

Eosinophils play an important role against parasitic infection, and produce antiviral molecules against respiratory viruses, including respiratory syncytial virus and influenza^23^. However, the role of eosinophils in SARS-Cov-2 infection is largely unknown. In this study, COVID-19 patients with eosinopenia were older than non-eosinopenia patients. Remarkably, Common symptoms included fever, cough, sputum, and fatigue were more frequent in eosinopenia patients. Meanwhile, eosinopenia patients had high proportion of dyspnea, gastrointestinal symptoms, and comorbidities. In additon, eosinopenia patients had lower SpO2, high AST levels and less counts of lymphocyte than non-eosinopenia patients. In our unpublished data, eosinopenia is more frequent than lymphopenia in COVID-19, implicating that eosnophils may be a better sensitive indicator than lymphocyte for evaluating the severity of COVID-19. In multivariate analysis, we found that less lymphocyte counts was independent risk factor of eosinopenia in COVID-19 patients. Apart from SARS-CoV-2, infection of MERS-CoV also had low counts of eosnophils enhancing the ability to detect patients with MERS-CoV^24^. Interestingly, effective administration of MERS-CoV vaccine could increase IL-5 and IL-13 cytokines leading to lung eosinophils elevated^25^. Coronavirus vaccines included SARS-CoV and MERS-CoV can develop lung eosinophilic immunopathology^26^. Additionally, eosinopenia is common in patients with H1N1 influenza^27^. Taken together, eosinopenia represents a novel phenotype in COVID-19 and has different characteristics from non-eosinopenia group.

Corticosteroids therapy is still controversial in the management of COVID-19. Early corticosteroids therapy is close associated with high blood SARS-Cov load and delayed MERS-Cov clearance^28^. Therefore, corticosteroids therepy should be avoided in patients with COVID-19 unless there are indications for moderate or severe ARDS, and septic shock. Short duration and low dose of corticosteroids therapy may be beneficial in management of severe COVID-19 patients ^28^. In this study, 100% of patients received antriviral treatment, and 50% of eosinopenia patients received corticosteroids therapy for most of eosinopenia patients were severe. One severe patient with eosinopenia accepted the treatments of convalescent plasma from recovered donors and tocilizumab. Duan K, et al provided evidence that convalescent plasma therapy could improve the clinical outcomes of severe COVID-19 patients^29^. Tocilizumab may be another effective treatment in COVID-19 patients with elevated IL-6 cytokine^30^. No fatal case was reported in our study, all COVID-19 patients with severe type were cured. The counts of eosnophils as well as lymphocyte were dramatically elevated and normalized in recovery patients with severe type compared with those at adimission during follow up.

There were some limitations in this study. First, despite this was a multi-center retrospective study in Anhui province, China, samples of this study were still small, which may lead to unavoidable bias. Second, clinical significance of eosinopenia in COVID-19 needs further investigations. Finally, this study was a observational study, more experimental research should be performed to reveal the pathogenesis of eosinopenia in COVID-19.

## Interpretation

Eosinopenia are very common in COVID-19 patient, particularly in severe patients, which may indicate a poor prognosis of COVID-19 patients. Common symptoms included fever, cough, sputum, and fatigue are more frequent in eosinopenia patients. In additon, eosinopenia patients had lower SpO2, high AST levels and less counts of lymphocyte. Eosinopenia may represent a novel phenotype in COVID-19.

## Data Availability

CY, ZY, ZM, ZL, LZ, DZ, YJ and YG contributed to the concept and design, analysis and interpretation as well as manuscript drafting. All authors read and approved the final manuscript.

## Guarantor

Yusheng Cheng takes responsibility for the content of the manuscript, including the data and analysis.

